# Structural Maintenance of Chromosomes 5/6 complex dysfunction enables tumor mutagenesis

**DOI:** 10.64898/2025.12.04.25341651

**Authors:** Thi Tran, Jiayi Fan, Xiaolan Zhao, Abby M. Green

## Abstract

The Structural Maintenance of Chromosomes (SMC) 5/6 complex is highly conserved and essential for mammalian development. SMC5/6 dysfunction in cells, model organisms, and patients with germline variants results in genome instability, though the exact function of the complex in genome maintenance remains enigmatic. Despite the importance of SMC5/6 in maintaining genome stability, the prevalence and consequences of somatic inactivation of SMC5/6 in cancer is understudied. Here we report a pan-cancer analysis of SMC5/6 dysfunction in cancer across three large databases. We identified thousands of tumors across all tissue types with copy number alteration and/or small variants in SMC5/6 genes. We found that deleterious variants in SMC5/6, but not copy number alterations, are associated with elevated tumor mutational burden (TMB). Mutagenesis in tumors with SMC5/6 variants was caused by polymerase epsilon (POLE) dysfunction and mismatch repair deficiency (MMRd). Patients in which SMC5/6 gene variants occur in tumor genomes exhibited improved survival relative to patients with non-altered SMC5/6 genes. This survival benefit was explained in part by a superior response to immunotherapy in a cohort of patients with colorectal cancer. Our findings demonstrate that dysfunction of SMC5/6 is predictive of elevated TMB and susceptibility to immunotherapy, indicating that SMC5/6 gene status should be considered in cancer diagnostic studies for prognostic implications and tailored therapeutics.

## INTRODUCTION

The three eukaryotic Structural Maintenance of Chromosomes (SMC) complexes, condensin, cohesin, and SMC5/6, are highly conserved and play important roles in genome integrity^1^. The complexes are structurally similar, comprised of a dimer of two distinct SMC proteins that can form elongated rings or filamentous structures, as well as accessory proteins^1^. SMC5/6 is distinct in that it contains protein-modifying subunits. Two of its non-SMC element (NSMCE) accessory subunits include the SUMO ligase (E3), NSMCE2, and a putative ubiquitin ligase, NSMCE1^2,3^. While cohesin and condensin are well known to organize chromatin by generating DNA loops^4,5^ (reviewed in *Ref 4 and 5* and references cited therein), the role of SMC5/6 appears to be more diverse and remains enigmatic^4,6^. The human SMC5/6 is comprised of six interacting proteins (SMC5, SMC6, NSMCE1-4) as well as the SLF1 and SLF2 subunits. Human SMC5/6 is best defined for for its role during interphase to aid in recombinational repair, replication completion, and coping with replication stress^7–12^. These functions are also assigned to the yeast Smc5/6, which can interact with double-stranded and single-stranded DNA gaps and generate DNA loops *in vitro*^13–23^.

A key to understanding the function of mammalian SMC5/6 is the recent reporting of several patients with germline SMC5/6 gene alterations associated with severe developmental diseases. Four studies over the last decade have detailed patients with biallelic mutations in SMC5^24,25^, NSMCE2^26^, NSMCE3^27^, and the chaperone SLF2^24^. All patients had microcephaly and developmental defects with some manifesting additional phenotypes including severe lung disease, dwarfism, insulin resistance, and bone marrow failure. While some variability exists among the syndromes reported, the patients displayed DNA damage and genome instability as evidenced by abnormal chromosomes, micronuclei, and nuclear bridges^24–27^. In comparison with these established effects of germline SMC5/6 dysfunction, the effect of SMC5/6 dysfunction in cancer is still an open question.

In a pan-cancer study, SMC5 and SMC6 were found to be among the top 50 most frequently mutated DNA damage genes^28^. Despite this frequency, a recent evaluation of SMC5/6 gene alterations in breast cancer is the only published study linking somatic dysfunction of the complex to tumor biology^29^. In breast cancer data collected from cBioPortal, alteration of copy number or DNA sequence in any SMC5/6 gene was associated with an increased aneuploidy score and poor overall survival^29^. With this link as a foundation, we sought to define the effect of somatic SMC5/6 dysfunction across human cancers.

We undertook an *in silico* study to define the frequency and impact of SMC5/6 dysfunction in human cancer. We examined three large datasets of annotated cancer genomes, namely The Cancer Genome Atlas (TCGA), Pan-Cancer Analysis of Whole Genomes (PCAWG), Colorectal Cancer Whole Genome Sequencing (Hartwig Medical Foundation). We found that a large fraction of tumors have alterations in SMC5/6 complex genes, either through copy number changes or small sequence variants (base substitutions, insertions/deletions). We found few “hotspot” or recurrent small variants, rather many different mutations throughout each of the nine SMC5/6 genes, consistent with a pattern evident in other cancer-related complexes. Small variants (hereafter denoted SmV), in comparison to copy number alteration, were associated with increased tumor mutational burden and mutagenesis caused by polymerase epsilon (POLE) defects and mismatch repair deficiency (MMRd). Importantly, SmV in SMC5/6 are associated with improved responses to immunotherapy. These data demonstrate the impact of SMC5/6 dysfunction in cancer genomes, a strong association between SMC5/6 dysfunction and POLE/MMRd-mediated mutagenesis, and implications for the potential to use SMC5/6 status as a biomarker for response to immune checkpoint blockade.

## RESULTS

### SMC5/6 variants occur across human tumors

To assess whether SMC5/6 dysfunction occurs in human cancer, we first analyzed all primary tumor exome sequences within TCGA. We investigated nine genes including those encoding for the six subunits of the core complex (SMC5, SMC6, NSMCE1-4), with two genes encoding NSMCE4 (NSMCE4A and NSMCE4B), and the SLF1 and SLF2 subunits (**Fig 1a**). In a previous study of SMC5/6 variants in cancer, all SMC5/6 gene aberrations were assessed in aggregate^29^. To discern whether abnormal gene products had a different impact than copy number variations, we generated two separate groups of human tumors: the first included somatic mutations (SmV including base substitutions and insertions/deletions) in SMC5/6 genes, and the second with copy number alterations (CNA) in SMC5/6 genes. The tumors with SmV in SMC5/6 genes were further divided into two subgroups: those predicted to be deleterious (i.e. frame shift, missense)^30,31^ versus those predicted to be benign (synonymous or non-synonymous non-deleterious (NS/ND) SmV,). Similarly, tumors with SMC5/6 copy number changes were categorized as low (<2) or high (>2) subgroups.

**Figure 1.**
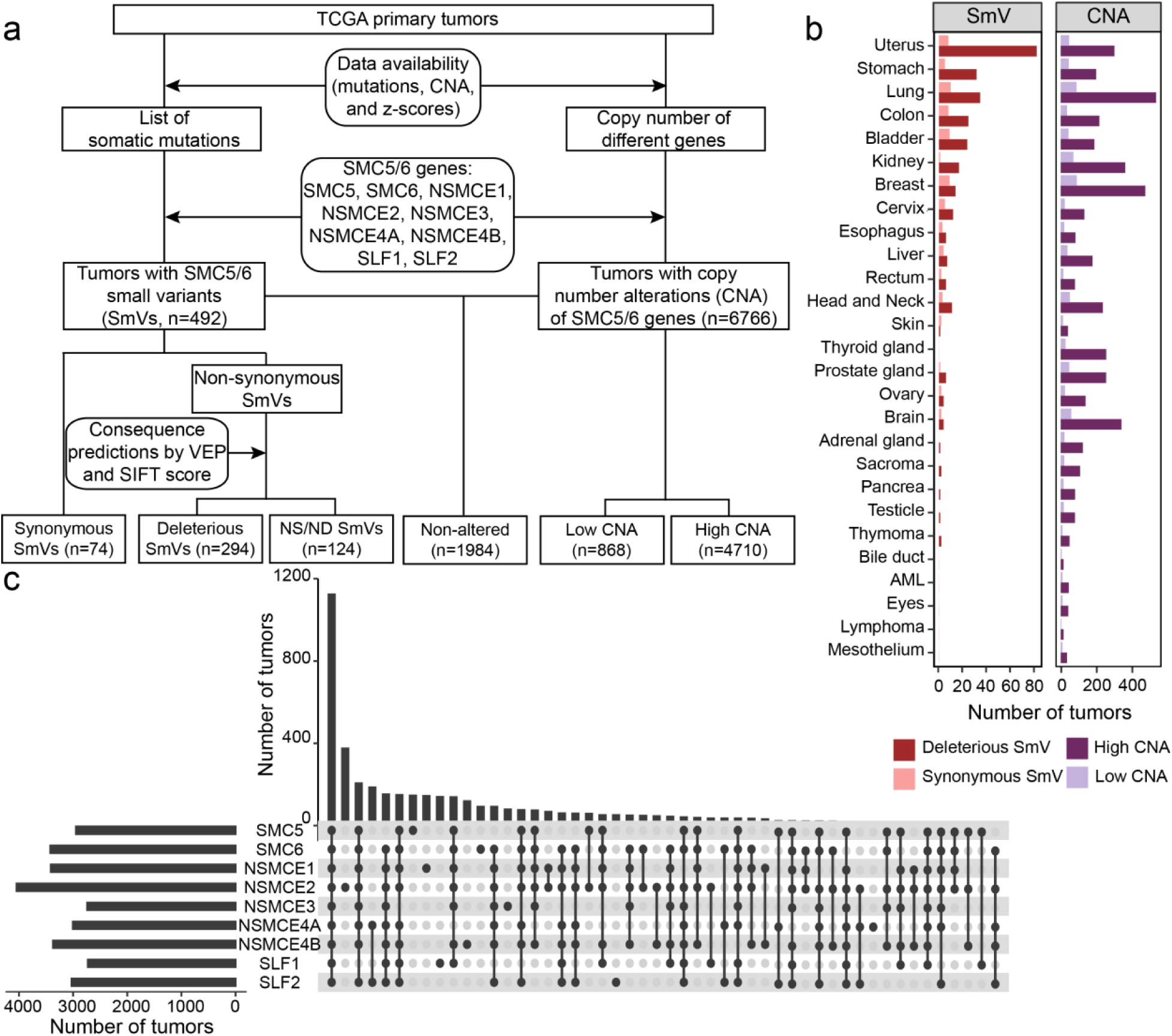
Alterations in SMC5/6 complex genes are evident throughout human cancers in TCGA. (**a**) Flow chart depicting the workflow to assess SMC5/6 gene alterations in tumor exome sequences from The Cancer Genome Atlas (TCGA). Exome sequences were filtered as shown. Small variants (SmV) include base substitutions and indels <50bp. Deleterious SmV were classified based on genetic (non-synonymous), evolutionary conservation (SIFT score), and biochemical (VEP) prediction methods. NS/ND SmV represent non-synonymous variants that were not predicted to be deleterious. Tumors with both SmV and CNA were included in both cohorts. Tumors with both high and low CNA of different SMC5/6 genes were excluded from all CNA cohorts. (**b**) Number of tumors with SMC5/6 gene alterations classified by tumor/tissue type. (**c**) UpSet plot depicting frequency of alterations (SmV and CNA) in SMC5/6 genes and combinations thereof.

Nearly three-quarters of tumors in TCGA displayed copy number alterations in SMC5/6 genes (n=6,766; of which 1,188 were excluded from further analysis for having both low and high CNA of different SMC5/6 genes), while 1,984 tumors retained wild-type SMC5/6 genes with a normal copy number. 492 tumors exhibited SmV in SMC5/6 genes, most of which were predicted to be deleterious (**Fig 1a**). The types of tumors exhibiting the most SMC5/6 gene SmVs (uterine, GI, lung) differed from those with the most SMC5/6 CNA (lung, breast, kidney, **Fig 1b**). The majority of tumors with alterations in SMC5/6 genes exhibited SmV and/or CNA of all nine genes (n=1,128, **Fig 1c**), likely reflecting global aneuploidy of a tumor. Tumors in which only one SMC5/6 gene was altered ranged from n=21-371 (**Fig 1c**). Among tumors with only a single SMC5/6 gene altered, *NSMCE2* was most frequently altered when assessing all possible gene alterations (**Fig 1c**), but *SMC6* was the most frequently altered when assessing only SmV (**Fig S1a**).

When SmV were assessed independently, the majority were found in *SMC5* and *SMC6* which comprise the largest open reading frames of all nine genes (**Fig S1a**). In contrast to the analysis of all variants (**Fig 1c**), little overlap across SMC5/6 genes was observed with respect to SmV (**Fig S1a**). Prior studies have demonstrated that loss of a single SMC5/6 complex component led to disruption of the entire complex and depletion of additional components^7,9,32^, thus the lack of overlapping gene SmVs in tumors may be due to the fact that disruption of a single SMC5/6 gene by SmV may interfere with function of the whole complex. To determine whether deleterious SmV occurred in a specific subunit, we quantified the number of tumors with SmV in each subunit. While tumors with SmV in *SMC5* and *SMC6* genes represented the largest group (**Fig S1b**), this is likely because they are the largest genes. When controlling for gene size, there did not appear to be a specific subunit that incurred more deleterious SmV (**Fig S1b**). Additionally, allele frequency of SmV was similar for all genes and indicated a pattern of allelically imbalanced SMC5/6 gene alteration (**Fig S1c**).

### Recurrent SMC5/6 SmV are rare

SMC5/6 is a multi-functional complex with both structural and enzymatic functions. Enzymatic functions can be mapped to specific domains of individual subunits and structural functions are enacted by multi-subunit interactions^2,3,33,34^. For example, structural studies of the yeast Smc5/6 complex defined dsDNA binding is enacted through the heads domains of Smc5 and Smc6, as well as on the Nse3 (NSMCE3) and Nse4 (NSMCE4) subunits^33,35^. We mapped the deleterious SmVs of SMC5/6 found in human tumors in TCGA to assess whether a specific domain was recurrently mutated. Domain annotations were based on integrating human SMC5/6 subunit domain mapping and the yeast Smc5/6 subunit structures mapped by cryo-EM studies^33,36,37^. We found that SMC5/6 SmV occurred throughout structural and functional domains (**Fig 2a**). Despite identifying 363 unique deleterious SmV in SMC5/6 genes across 492 human tumors, we found very few recurrent mutations (**Fig 2a**). We assessed the SmV that occurred most frequently in each SMC5/6 gene and found approximately 2/3 were missense mutations (**Fig 2b**). When these variants were analyzed by AlphaFold missense, only 50% had a pathogenicity score >0.5 (**Fig 2b**). To understand the pathogenic effects of frequent SmVs, we assessed both associated tumor mutational burden (TMB) and homology to yeast Smc5/6 structure. We found that the mutational burden within tumors with most frequent SmV was substantially higher than the average TMB for all tumors with SMC5/6 deleterious SmV (**Fig 2c**), suggesting that disruption of SMC5/6 through frequently occurring variants is associated with higher levels of tumor mutagenesis.

**Figure 2.**
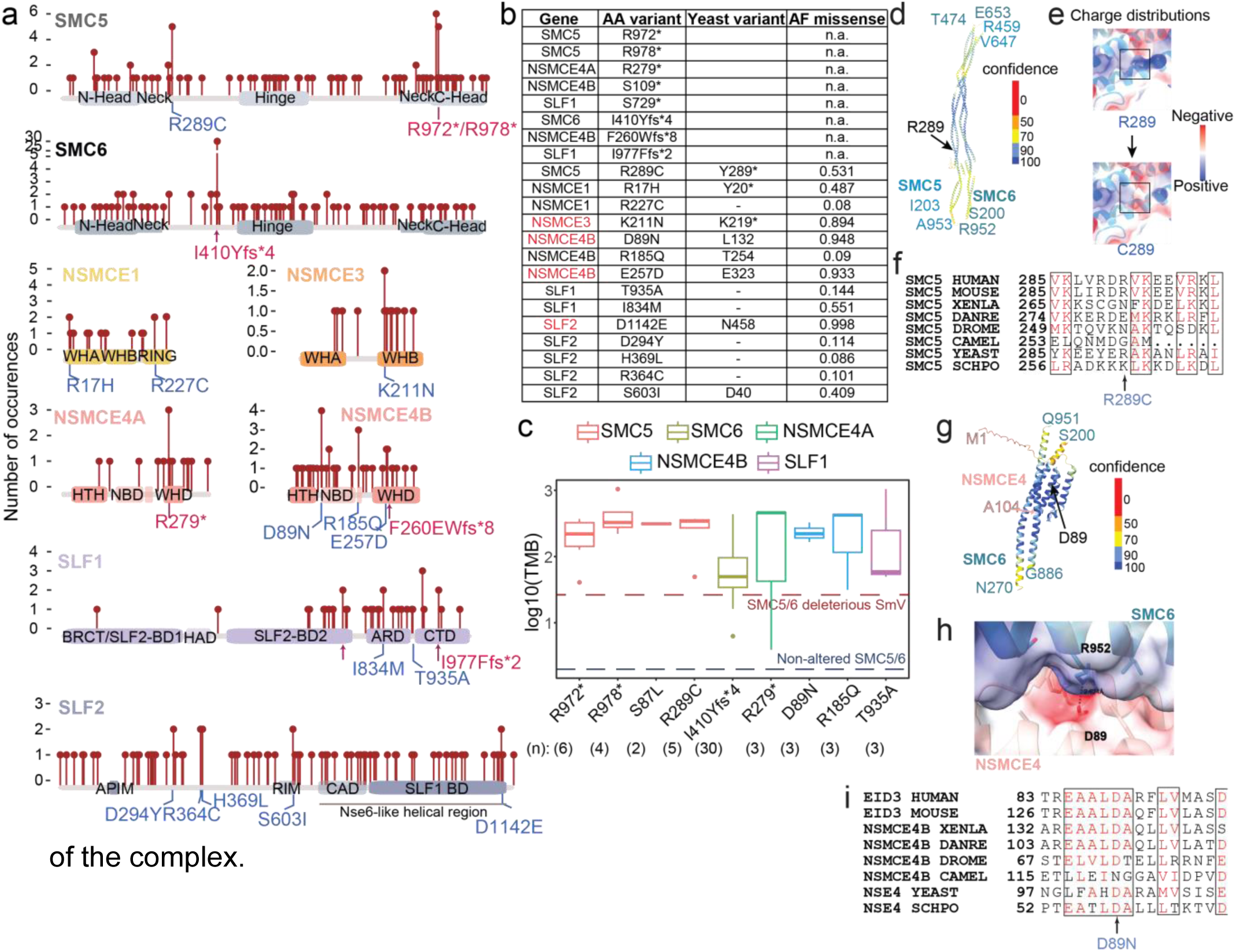
SMC5/6 dysfunction in cancer is not caused by hotspot mutations. (**a**) Lollipop chart showing location and frequency of deleterious SmV detected in TCGA tumor exomes within SMC5/6 complex genes. Each SMC5/6 subunit is shown with its known conserved structural and functional domains annotated. Two types of mutations are highlighted: truncating mutations (light blue) and single amino acid substitutions (dark blue). (**b**) Table depicts most frequently occurring mutations for each SMC5/6 gene including amino acid (AA) change, yeast homology site, and AlphaFold missense pathogenicity score. Homologous sites in yeast are not listed for poorly conserved or unstructured regions. (**c**) Box and whisker plot shows tumor mutational burden (TMB) in exomes containing most frequently occurring SMC5/6 gene SmV. Red dotted line shows average TMB for exomes with deleterious SMC5/6 SmV, blue dotted line shows average TMB for exomes with no SMC5/6 gene alteration. Number in parenthesis indicates the number of times each mutation was detected in independent exomes. (**d**) AlphaFold-predicted model of the human SMC5/6 arm region (ipTM=0.52, pTM=0.52), colored by model confidence (pLDDT). The residue ranges used for prediction are indicated in the corresponding subunit colors, and the mutated site is marked in black. (**e**) Close-up view of residue R289 in SMC5. Substitution of arginine (R) with cysteine (C) reduces local positive charge and may disrupt electrostatic interactions along the coiled-coil interface, potentially affecting local stability or intrasubunit contacts. (**f**) Sequence alignment of the SMC5 arm region across representative species shows limited conservation at position 289, suggesting that this residue may contribute to fine-tuning rather than an essential conserved function within the SMC5 arm. (**g**) AlphaFold-predicted model of the human EID3 helix-turn-helix (HTH) domain and its interface with part of SMC6 arm (ipTM=0.83, pTM=0.78). The predicted model is colored by pLDDT. The residue range used for prediction is indicated in subunits’ corresponding colors, and the mutated site is indicated in black. (**h**) A close-up view of D89 with electrostatic surface and alignment of NSMCE4 D89 residue. NSMCE4-D89 is predicted to interact with SMC6-R952. Such interaction is also seen in the yeast Smc5/6 structure (Yu et al, 2022, PDB: 7TVE). (**i**) Sequence alignment shows that D89 is relatively well conserved. WH: Winged-Helix; HTH: Helix-Turn-Helix. BRCT: Breast Cancer gene C-terminal; BD: binding domain; IDR: intrinsic disordered region; ARD: Ankyrin Repeat Domain; CTD: C-Terminal domain.

As no cryo-EM map of the human SMC5/6 complex is available, we relied on the structure of the homologous yeast Smc5/6 complex^33,36,37^ and AlphaFold predicted structures to assess the functional impact of SMC5/6 variants found in cancer. While not all tumor-associated SMC5/6 SmV had homologous sites in yeast (**Fig 2b**), two recurrent missense mutations, SMC5 R289C and NSMCE4B D89N, were mapped to homologous regions and subjected to homology modeling. R289C lies in the coiled-coil domain of the SMC5 protein (**Fig 2d**) and is predicted to reduce local positive charge, which may disrupt electrostatic interactions along the coiled-coil interface between SMC5 and SMC6, potentially affecting stability or intrasubunit contacts (**Fig 2e**). Sequence alignment of the SMC5 arm region across species shows limited conservation at position 289 (**Fig 2f**), suggesting that the residue may contribute to fine-tuning rather than an essential conserved function. NSMCE4B D89 lies in the helix-turn-helix (HTH) domain of the protein and its interface with part of the SMC6 arm region (**Fig 2g**). NSMCE4B D89 is relatively well conserved and is predicted to interact with SMC6 R852 (**Fig 2h-i**), an interaction that is also seen in the yeast Smc5/6 structure^33^. These predictions suggest that cancer-associated SMC5/6 variants have the potential to impact integrity and function.

When mapped to lengthwise renderings of SMC5/6 genes, the pattern of SmV appeared to be “long and low” in striking contrast to that of well-studied oncogenes and tumor suppressors such as TP53 and PIK3CA which exhibit specific “hotspot” cancer-associated mutations (**Fig S2**). Interestingly, this long and low pattern of SmV was also observed for cohesin gene mutations in cancer (**Fig S3**). Cohesin is an SMC protein complex that, in contrast to SMC5/6, has an established role in tumorigenesis^38^. These data suggest that gene variants may occur throughout protein complexes in cancer. Given the essential interactions between many genes and domains of a protein complex, a long and low pattern of variants may be equally disruptive to that of hotspot mutations in individual oncogenes.

### SMC5/6 dysfunction is associated with increased tumor mutational burden

To assess the functional impact of SMC5/6 dysfunction, we evaluated genome instability in cancers with altered SMC5/6 genes in TCGA. When tumors with SMC5/6 SmV and CNA were pooled, we found an increase in TMB relative to SMC5/6 WT tumors (**Fig 3a**). Interestingly, we found no difference in aneuploidy between these groups (**Fig 3a**), differing from a recently reported association between SMC5/6 and aneuploid tumor genomes^29^. Based on these data, we asked whether SMC5/6 SmV and CNA similarly affected TMB. A slight increase in TMB was found in the pool of tumors with SMC5/6 CNA relative to WT, however a substantial increase in TMB in tumors with SMC5/6 SmV relative to tumors with either SMC5/6 CNA or non-altered SMC5/6 was detected (**Fig 3b**). When copy number alterations in SMC5/6 genes were further narrowed to groups of low (<2 copies) or high (>2 copies), we found similar levels of TMB when compared to tumors with non-altered SMC5/6 genes (**Fig S4a-b**). When comparing tumors grouped by SmV, CNA, or non-altered SMC5/6 genes, we again found no alteration in aneuploidy score (**Fig 3b, Fig S4b**).

**Figure 3.**
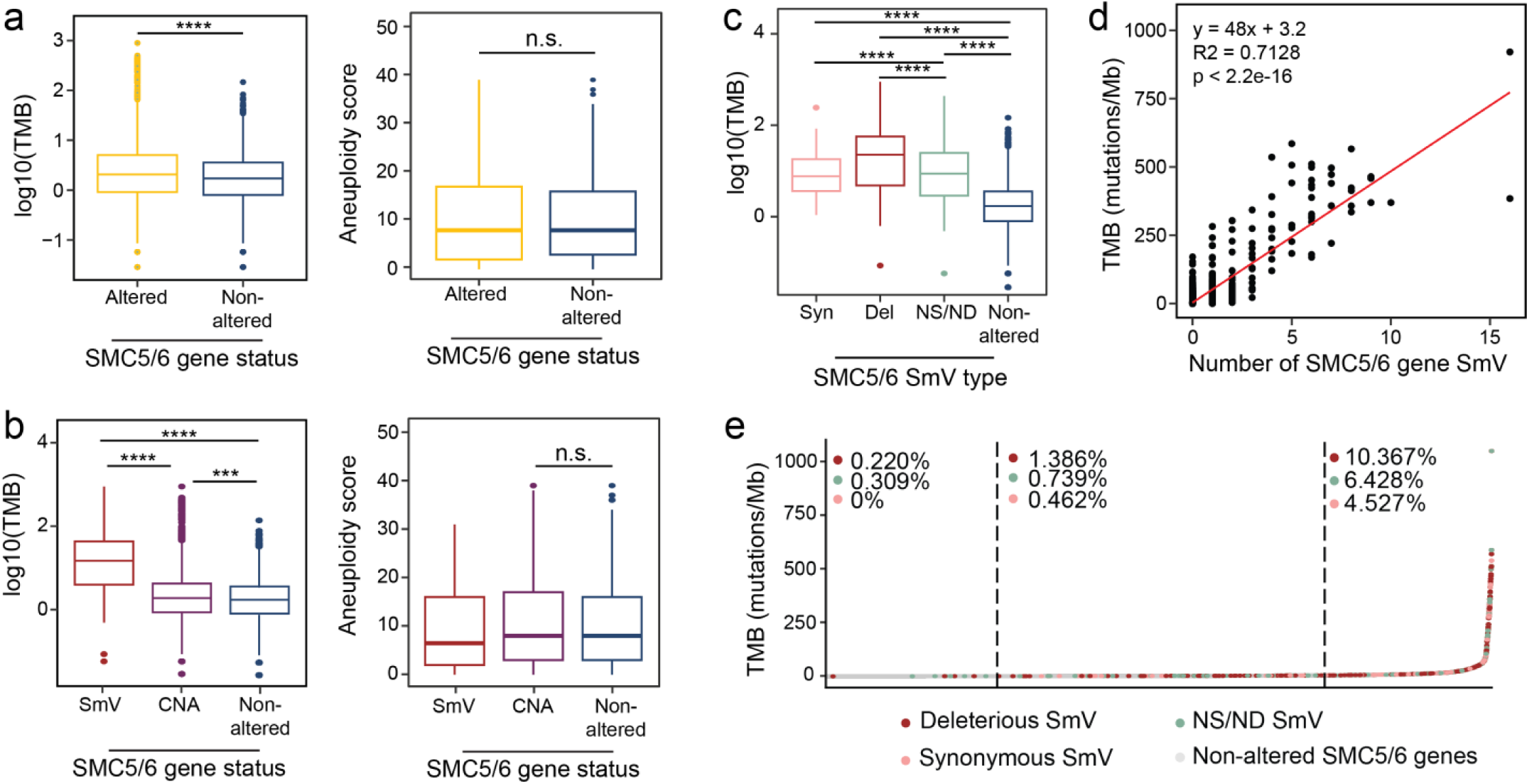
SMC5/6 dysfunction correlates with elevated tumor mutational burden. (**a-b**) TMB and aneuploidy score of TCGA exomes (**a**) with or without any alteration (SmV or CNA), or (**b**) divided further into SmV, CNA, or non-altered SMC5/6 genes. (c) TMB of TCGA exomes categorized by type of SMC5/6 SmV. Significance determined by Willcoxon rank-sum test; **** p<0.0001. (**d**) Dot plot depicting correlation between TMB and number of SMC5/6 gene SmV per exome. A linear best-fit regression is shown in red. The corresponding linear equation, Pearson correlation coefficient (R^2^), and p-value are provided. (**e**) TMB of each exome in TCGA is plotted from lowest to highest. Color designates type of SMC5/6 SmV in each exome. Dotted lines denote the lower and upper quartiles. Frequency of each type of SmV per section is shown. X^2^ test for independence was 948.59, df = 6, p < 0.0001.

To complement the analysis of TCGA exomes, we analyzed tumor whole genomes from PCAWG and found 1,923 tumors with SMC5/6 gene alterations (**Fig S5a**). Similar to tumors in TCGA, those in PCAWG with SMC5/6 SmV displayed elevated tumor mutational burden relative to tumors with non-altered SMC5/6 genes (**Fig S5b**). While PCAWG tumors with SMC5/6 CNA had an elevated TMB relative to those with non-altered SMC5/6, the level remained significantly lower than tumors with SMC5/6 SmV (**Fig S5b**). These data indicate that SMC5/6 disruption, and specifically gene variants, are associated with increased tumor mutational burden.

When assessed independently, variants in all SMC5/6 genes were associated with elevated TMB in TCGA exomes (**Fig S6a**). Similarly, CNA of each SMC5/6 gene assessed independently did not reveal any association with altered TMB (**Fig S6b**). However, when we categorized tumors by the type of SmV – deleterious, synonymous, or neither – we found that SmV predicted to be deleterious to SMC5/6 gene products were associated with the highest mutational burden (**Fig 3c**). Further, a striking correlation between number of SMC5/6 SmV within a tumor and mutational burden was observed (**Fig 3d**). These data suggest that the degree of SMC5/6 dysfunction incurred by a tumor may impact mutational burden.

### Elevated TMB and SMC5/6 SmV: the chicken or the egg?

The correlation between SMC5/6 SmV and high TMB can be explained by two models: the first model suggests that dysfunction of SMC5/6 caused by SmV may enable an increase in TMB, while the second model posits that tumors with a high mutational rate could incidentally result in SmV of SMC5/6 genes. The second model, therefore, predicts an equal distribution of variant types across each SMC5/6 gene. Instead, we found that the majority of SmV were predicted to be deleterious in nearly all SMC5/6 genes (**Fig S1b**). The second model also predicts a similarly elevated TMB across all SmV types. In contrast, we found that deleterious SmV are associated with significantly higher TMB than synonymous or NS/ND variants (**Fig 3c**). In this analysis, we sorted TCGA exomes from lowest to highest TMB, divided the cohort into quartiles, and assessed the occurrence of SMC5/6 SmV in each quartile (**Fig 3e**). The second model predicts that the type of SmV would occur with similar frequency across each quartile. While deleterious, synonymous, and NS/ND SmV were all enriched in highest quartile, the fraction of tumors with deleterious SmV was significantly higher than those with synonymous or NS/ND SmV (*X*^2^ (6, n=8,809)= 948.59, *p*<0.0001). These data support the first model of directional correlation in which dysfunction of SMC5/6 drives TMB.

### SMC5/6 dysfunction predicts response to immunotherapy

To assess the phenotypic effect of SMC5/6 dysfunction in cancer, we evaluated overall survival of patients with non-altered or altered SMC5/6. From tumor genomes in TCGA, we found that any alteration in SMC5/6 was associated with improved survival (**Fig 4a**). Tumors with SMC5/6 SmV had improved survival over those with other SMC5/6 alterations (**Fig 5b**), and deleterious SMC5/6 SmV predict the best survival among all tumors stratified by SMC5/6 status (**Fig 5c**). Tumors with high TMB are often sensitive to immunotherapeutic approaches^39,40^. The “hypermutator” tumors are more susceptible to immune clearance, therefore more sensitive to immune checkpoint blockade to enhance tumor killing by cytotoxic T-cells. We hypothesized that the improvement in survival of patients with tumors harboring SMC5/6 SmV may be due to susceptibility to immunotherapy. To test this possibility, we interrogated a dataset of metastatic colorectal cancer from the Hartwig Medical Foundation with thorough annotation of treatment and survival^41^. A subgroup analysis of colorectal tumors with SMC5/6 SmV demonstrated improved survival in those patients treated with immunotherapy relative to other treatment groups (**Fig 4d**). We assessed patient survival following specific therapies and found patients with SMC5/6 SmV demonstrated improved survival upon treatment with immunotherapy as compared to patients without SMC5/6 SmV (**Fig 4e**). From these findings across two large databases of cancer, we conclude that SMC5/6 SmV improves responses to immunotherapy, likely through increasing tumor mutational burden.

**Figure 4.**
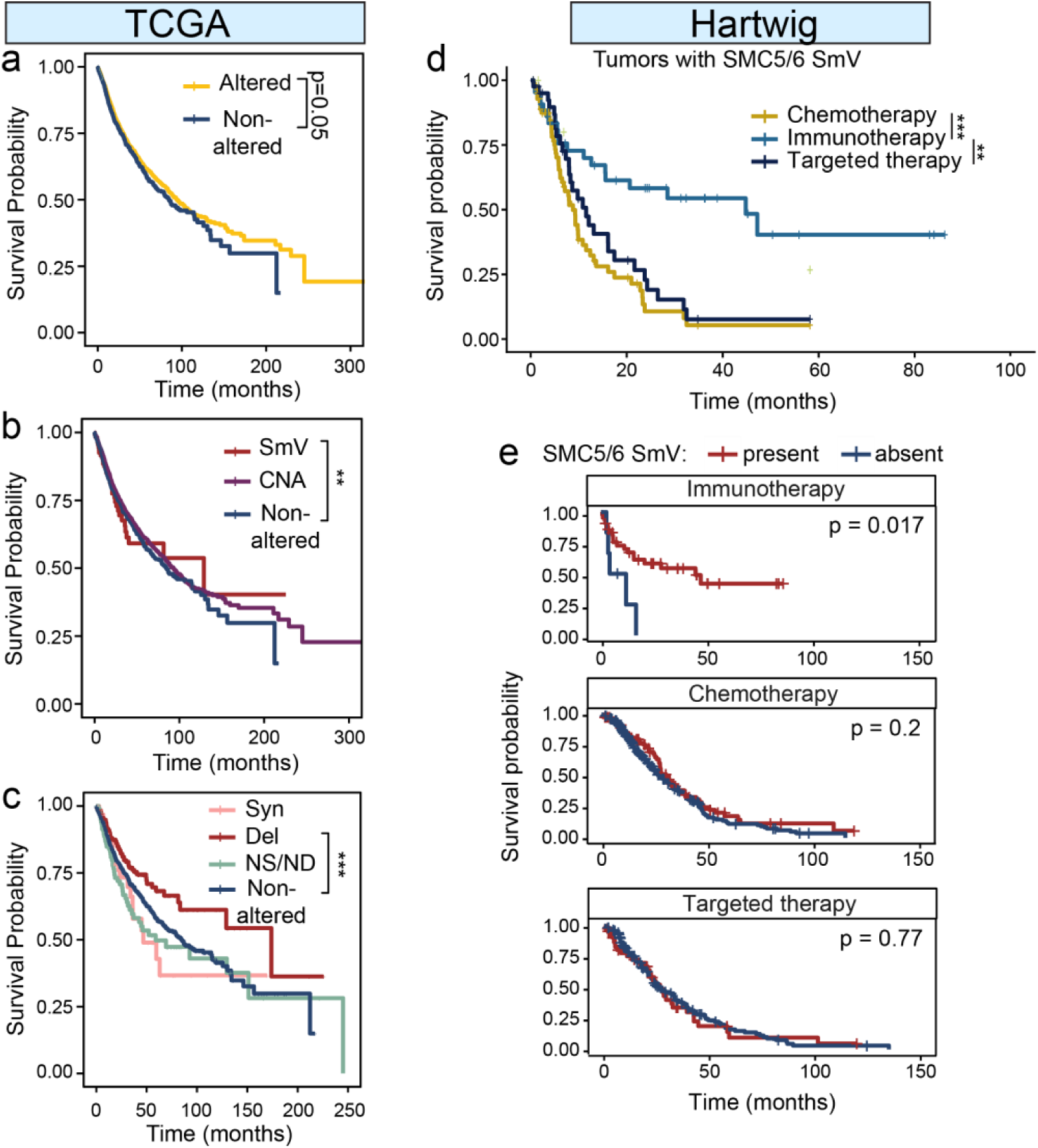
SMC5/6 dysfunction is correlated with improved patient survival. (**a-c**) Kaplan-Meier curves show survival of patients in TCGA cohort with (**a**) any SMC5/6 gene alteration or non-altered SMC5/6 genes, (**b**) SmV, CNA, or non-altered SMC5/6 genes, (**c**) categorized by type of SMC5/6 SmV. (**d-e**) Kaplan-Meier curves show survival of patients in Hartwig colorectal cancer cohort separated by therapy received. (**d**) All patients with SMC5/6 SmV detected in tumor genomes are shown according to treatment type. (**e**) Recipients of specific cancer therapies are stratified by SMC5/6 SmV status (present v. absent). P-values determined by log-rank test.

**Figure 5.**
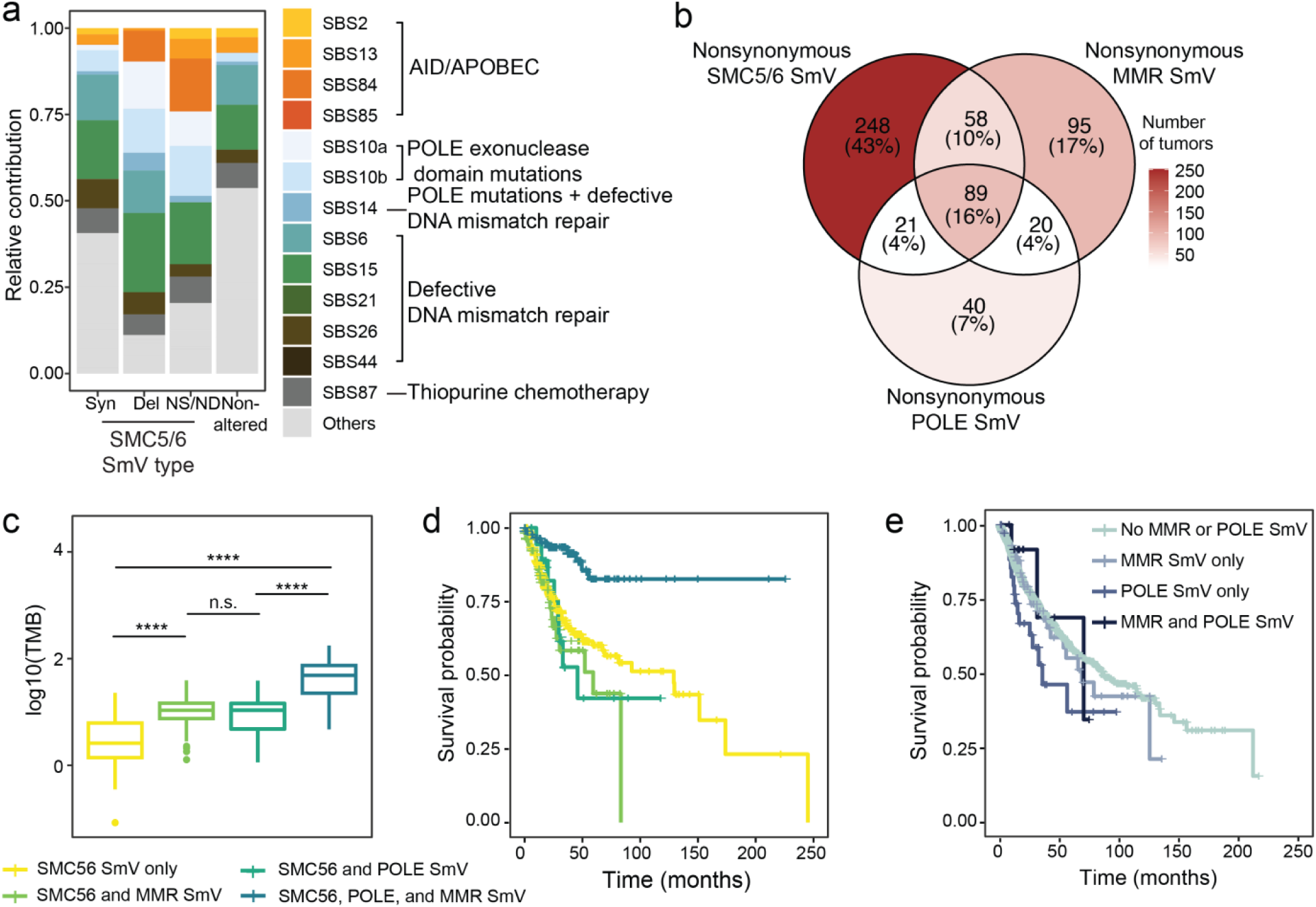
Increased TMB in SMC5/6-variant tumors reflects POLE dysfunction and MMR deficiency. (**a**) Relative contribution of single base substitution (SBS) signatures from TCGA exomes categorized by SMC5/6 SmV type. Colored signatures are designated with etiology in legend; these signatures comprised >5% relative contribution of TMB in SMC5/6 deleterious SmV cohort. Signatures in gray are designated “others” and comprised <5% relative contribution of TMB in deleterious SmV cohort. (**b**) Venn diagram shows confluence of TCGA tumors with non-synonymous SmV in SMC5/6 genes, mismatch repair (MMR) genes, and polymerase epsilon (POLE). Color gradient shows number of tumors in each group. (**c**) TMB and (**d**) Kaplan-Meier curve of TCGA exomes with non-synonymous SmV in SMC5/6 genes, MMR genes, and/or POLE. Significance determined by Wilcoxon rank-sum test ****p<0.0001, pairwise comparisons between the cohort with SmV in all (SMC5/6, MMR, POLE) relative to each other group is shown. (**e**) Kaplan-Meier curve of TCGA exomes with non-synonymous SmV in MMR genes, and/or POLE but non-altered SMC5/6 genes. All pairwise comparisons are not statistically significant.

### SMC5/6 dysfunction enables mutagenesis by PolE defects and MMRd

To define the mutational processes that drove elevated TMB in tumors with SMC5/6 variants, we pooled TCGA exomes based on SMC5/6 SmV type and assessed for single base substitution (SBS) mutational signatures. Within tumor exomes with SMC5/6 SmV, we found an enrichment in SBS signatures caused by polymerase epsilon (POLE) mutations and by defective mismatch repair (MMRd) indicating that these mutational processes are correlated with SMC5/6 dysfunction (**Fig 5a**). MMRd and POLE mutational processes are well-defined to generate a “hypermutator phenotype” in colorectal, endometrial, and other cancers^42–45^. To determine the effect of SMC5/6 dysfunction in combination with these mutagenic processes, we defined subgroups of TCGA tumors based on all nonsynonymous variants in SMC5/6 genes, MMR genes, and POLE genes, and combinations of these three subgroups (**Fig 5b**). As expected, the subgroups were enriched for SBS signatures defining each deficiency: MMR variant tumors were enriched for MMRd signatures and POLE variant tumors had elevated POLE-associated SBS signatures (**Fig S7a**). We found that SMC5/6 variants synergize with MMRd and POLE dysfunction, leading to elevated mutational burden and improved survival relative to tumors with only SMC5/6 variants (**Fig 5c-d**). Control tumors without variants in SMC5/6, MMR, or POLE genes or with variants in only one category displayed worse patient survival than those with combinations of variants (**Fig S7b-c**). The combination of all three (SMC5/6, MMR, and POLE variants) resulted in the highest TMB and best survival (**Fig 5c-d**). Interestingly, among exomes with non-altered SMC5/6 genes, nonsynonymous SmV in MMR genes and POLE did not result in better patient outcomes (**Fig 5e**). These data suggest that loss of SMC5/6 function in cancer exacerbates mutagenesis caused by POLE defects and MMRd, and may serve as a biomarker for response to immune checkpoint blockade.

## DISCUSSION

The SMC5/6 complex has multiple functions in genome maintenance and germline mutations in SMC5/6 genes are detrimental to the development and health of humans and model organisms. However, little is known about the consequences of somatic SMC5/6 inactivation in cancer. In this pan-cancer study, we show that disruption of SMC5/6 genes correlates with high TMB. Similarly elevated TMB is found in other tumors with DNA repair defects^46,47^. In principle, genome plasticity associated with SMC5/6 dysfunction could result in cancer cell evolution and heterogeneity, leading to poor patient outcomes^48^. Indeed, this was suggested in a study of SMC5/6 alterations in breast cancer^29^. In contrast, our data demonstrate that, across all cancers, SmV in SMC5/6 are associated with improved survival (**Fig 4a-c**). In a dataset of several hundred patients with metastatic colorectal cancer, SMC5/6 SmV correlated specifically with improved responses to immunotherapy (**Fig 4d-e**). These findings are analogous to the reported responses to immunotherapy in hypermutated tumors with MMRd and POLE dysfunction^39,40^. Interestingly, we find that somatic disruption of SMC5/6 is correlated with mutagenesis from MMRd and POLE dysfunction. Additionally, SMC5/6 variants in cancer augment responses to immunotherapy in tumors with MMRd or POLE mutations, resulting in superior patient survival relative to MMRd or POLE mutations alone (**Fig 5**). These data indicate that loss of SMC5/6 function in cancer may enable hypermutator phenotypes caused by MMRd and POLE dysfunction.

A prior relationship between Smc5/6 and POLE was established in yeast studies in which Smc5/6 was found to directly interact with and sumoylate the polymerase epsilon (pol ε) Pol2 subunit^49,50^. Sumoylation of Pol2, which occurred during S phase and upon replication stress, was necessary for replication progression and prevention of recombination-mediated chromosomal rearrangements^49,50^. In cancer, replication stress is ubiquitous, thus replication polymerases and replication stress responses are essential to maintain high rates of DNA synthesis, enable genome duplication, and promote cell and tumor growth. Though the presence and function of sumoylated pol ε in mammalian cells has not be confirmed, it is conceivable that dysfunction of SMC5/6 in cancer would limit pol ε sumoylation leading to lack of polymerase function and/or fidelity and an increased mutational rate. This model would explain the finding of POLE dysfunction mutational signatures in tumors with deleterious variants in SMC5/6. No correlation between SMC5/6 and MMR has been reported, though our data suggest that SMC5/6 supports MMR, a prediction warranting future biological studies.

The specific function or domain of the multi-functional SMC5/6 complex that supports high-fidelity DNA synthesis by pol ε and DNA repair by MMR is unknown since no hotspot SmVs in SMC5/6 were found in our study, similar to a previous report^29^. The “long and low” pattern of SMC5/6 SmV was also noted in cohesin genes in our study and in others in which expanded cohorts were analyzed^38^, but in stark contrast to most tumor suppressor and oncogene SmV. We postulate that somatic dysfunction of protein complexes can be achieved through numerous routes rather than a single point of inactivation or hyperactivation. This is supported by homology modeling of SMC5/6 cancer variants using the yeast Smc5/6 structure which predicted subtle changes to functional domains and protein subunit interactions. Most cancer-associated SMC5/6 SmV were hetero/hemizygous, which is expected given that SMC5/6 is essential for cell proliferation. We found a cohort of tumors that harbored more than one variant in SMC5/6 and were associated with particularly elevated TMB; it is possible that combinations of allelically imbalanced SMC5/6 SmV cause more substantial complex dysfunction. Our *in silico* study did not evaluate the specific function of SMC5/6 SmV (or combinations of SmV), but the correlation between somatic SMC5/6 dysfunction and increased TMB demonstrates a previously undefined role for SMC5/6 in preventing base substitutions.

Our study lays a foundation for further investigation into the clinical implications of SMC5/6 dysfunction in cancer. SMC5/6 variants correlate with a survival benefit in patients treated with immunotherapy and in cases of combined SMC5/6 variants with POLE mutation or MMRd. These findings suggest that SMC5/6 genes should be included in diagnostic targeted sequencing panels to enable prognostic predictions and tailored therapeutic approaches.

## METHODS

### SMC5/6 gene alteration identification and cohort analysis

Our study includes 9,614 samples from TCGA, 2,778 samples from PCAWG, and 724 samples from Hartwig Medical Foundation. Only primary samples from the TCGA (n=9614) and PCAWG (n=2778) projects were included in this study. Data obtained from the Hartwig Medical Foundation consisted of metastatic colorectal cancer genomes^41^. Samples missing data from somatic variant calling, copy number alterations, or survival information were eliminated from cohort analysis. The final dataset contained 8,809 TCGA samples, 2,583 PCAWG samples, and 545 Hartwig samples. Hartwig variant calling files were processed with VEP (release 112.0) and vcf2maf (https://github.com/mskcc/vcf2maf) to predict mutation consequences. Small variants (SmV) with either HIGH VEP impact^31^ or SIFT score < 0.05^30^ were predicted to be deleterious. Tumors were selected for exonic SmVs in at least one of the SMC5/6 genes (SMC5, SMC6, NSMCE1, NSMCE2, NSMCE3, NSMCE4A, NSMCE4B, SLF1, and SLF2), and classified as deleterious, synonymous, or nonsynonymous-nondeleterious (NS/ND). The sorting order for these cohorts were deleterious, NS/ND, and synonymous to account for samples with multiple types of SMC5/6 SmVs. Tumors were also selected for having abnormal copy number (≠ 2) of at least one of the SMC5/6 gene segments. For both TCGA and PCAWG data, low copy number variants (low CNV) cohort consisted of tumors with copy number less than 2, while high CNV cohort consisted of those with copy number greater than 2. With Hartwig data, low CNV group contained samples with maxCopyNumber ≤ 1.5, and high CNV group were made up of tumors with minCopyNumber ≥ 2.5. Samples with both low and high CNV of SMC5/6 genes were eliminated from later analysis. Samples with both SMC5/6 SmVs and abnormal copy number of SMC5/6 genes segments were included in both SmV and CNV groups. Samples with neither of these genetic alterations in any of the SMC5/6 genes were assigned to the non-altered group. As there were not many non-altered samples identified in the Hartwig dataset, abnormal CNV and non-altered cohorts were grouped together in later analysis (labelled as “SmV absent”).

### Co-occurrence of SMC5/6 alterations, SmV type and frequency, and variant allele frequency (VAF)

Tumors with multiple SMC5/6 alterations were identified. The sorting order (deleterious, NS/ND, and synonymous) was removed in this analysis to best reflect the number of tumors that carry each type of SMC5/6 SmV. Co-ocurrence of alterations (CNA and/or SmVs) across SMC5/6 genes was depicted using package UpSetR (version 1.4.0) in R. Mutation frequency was calculated using the equation below:

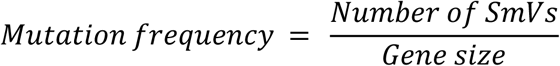

where number of SmVs was the total number of mutation events of a SMC5/6 gene identified in TCGA. VAF was calculated as

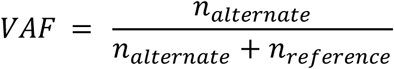

where *n*_*alternate*_ was the number of reads with the altered base, and *n*_*reference*_ was the number of reads with the reference base at the locus.

### SMC5/6 SmV mapping and yeast homology assessment

All deleterious SMC5/6 variants identified in TCGA were mapped by protein location and their resulted changes in amino acids sequence. The frequency of their occurrence in TCGA was then counted and visualized with lollipop chart generated using ggplot2 R package (version 4.0.0). Cancer-associated SMC5/6 SmV were aligned with each SMC5/6 subunit annotated with its known conserved structural and functional domains annotated. Deleterious truncating mutations and single amino acid substitutions were considered for homology analysis. Mutations located in unstructured regions were not further analyzed due to insufficient model reliability. AlphaFold was used to predicted models of the human SMC5/6 arm and EID3 helix-turn-helix (HTH) domain and its interface with part of SMC6 arm.

### Tumor mutational burden, aneuploidy, and mutational signature analysis

#### Tumor mutational burden (TMB) and aneuploidy score analysis

TMB (total number of mutations per Mb) was calculated as the number of all SmVs occurred in a tumor, divided by either 30 (for TCGA exomes) or 2,800 (for PCAWG and Hartwig genomes). Silent and intronic SmVs were included in this calculation. Pairwise comparisons of TMB between cohorts were conducted and visualized using R package ggpubr (version 0.6.0). The relationship between number of SMC5/6 SmVs in each tumor and corresponding TMB was visualized as a linear regression model and evaluated with Pearson correlation. To analyze the distribution of SMC5/6 SmVs with different predicted outcomes across the TMB spectrum, sorting bias (deleterious > NS/ND > synonymous) was removed. The distribution was evaluated with Chi-square test. TCGA aneuploidy scores and PCAWG ploidy scores were readily available in cBioPortal (see Data availability).

#### Single-base-substitution signatures (SBSsig) analysis

Trinucleotide context of each tumor was generated by counting the number of single base substitution events (C>T, C>A, C>G, T>C, T>A, T>G), along with their 5’ and 3’ adjacent base that the tumor harbored. The mutational contexts of different samples were then added together to form a single trinucleotide context matrix that represented the study cohorts to which they belonged. These matrices were refitted to COSMIC SBSsig (version 3.2) using R package MutationalPatterns (version 3.14.0). The result was a combination of established COSMIC SBSsig that best reconstructed the mutational matrix of each cohort.

#### Analysis of tumors with co-mutated SMC5/6, POLE, and MMR genes

TCGA exomes were selected as whether they had nonsynonymous SmV in at least one of the POLE (POLE, POLE2, POLE3, POLE4) or MMR (MLH1, MLH2, MLH3, MSH2, MSH6, PMS1, MS2) genes. Tumors were further categorized as to whether they contained nonsynonymous SmV or non-altered SMC5/6 genes.

### Survival analysis

Overall survival time and status of patients included in TCGA and PCAWG were obtained from cBioPortal (see Data Availability). Overall survival time for patients from the Hartwig dataset was counted from the day they joined the study to the date of their death or last follow up date. Only patient response to treatment admitted after biopsy was accessed in this study. Constructing and analyzing Kaplan-Meier curves was handled using R packages ggsurvfit (version 1.1.0), survminer (version 0.4.9), and survival (version 3.8.3).

### Statistical analysis

All statistical tests were performed in R with ggpubr (version 0.6.0). Pairwise comparison for TMB, aneuploidy, and ploidy score was accessed by Wilcoxon rank-sum test. Constructing and analyzing Kaplan-Meier curves was handled using R packages ggsurvfit (version 1.1.0), survminer (version 0.4.9), and survival (version 3.8.3). Pairwise log-rank test was used for survival analysis.

### Data availability

Somatic variant calls and copy number information of the TCGA projects were obtained using R package TCGA biolinks (version 2.32.0), while whole genome sequencing data of the ICGC was downloaded at https://dcc.icgc.org/releases/PCAWG (release 28). Clinical information, aneuploidy and ploidy scores for both TCGA and PCAWG (ICGC) dataset were obtained from cBioPortal on January 15^th^, 2024. Sample and patient identification numbers were utilized to match data across these databases. Somatic variant calling and clinical data of colorectal metastases were performed and reported by the Hartwig Medical Foundation (study number NCT01855477).

## ACKNOWLEDGEMENTS

We thank all members of the Green and Bednarski Labs for helpful discussions and input. We thank Ilham Matikar and Naser Salas-Husain for technical support. This study was funded by support from the DOD CA200867 (AMG), ACS RSG1313877 (AMG), NIH R01GM153923 (AMG), and Pedal the Cause.

## AUTHOR CONTRIBUTIONS

TT and AMG designed experiments. TT performed informatic experiments, analysis, interpretation and statistical tests. JF performed structural modeling experiments and interpretation. All authors conceptualized the study. XL and AMG supervised the study. AMG wrote the manuscript with input from all authors.

## Competing Financial Interests

No disclosures or conflicts of interests were reported.

## SUPPLEMENTARY DATA

**Supplemental Figure 1.**
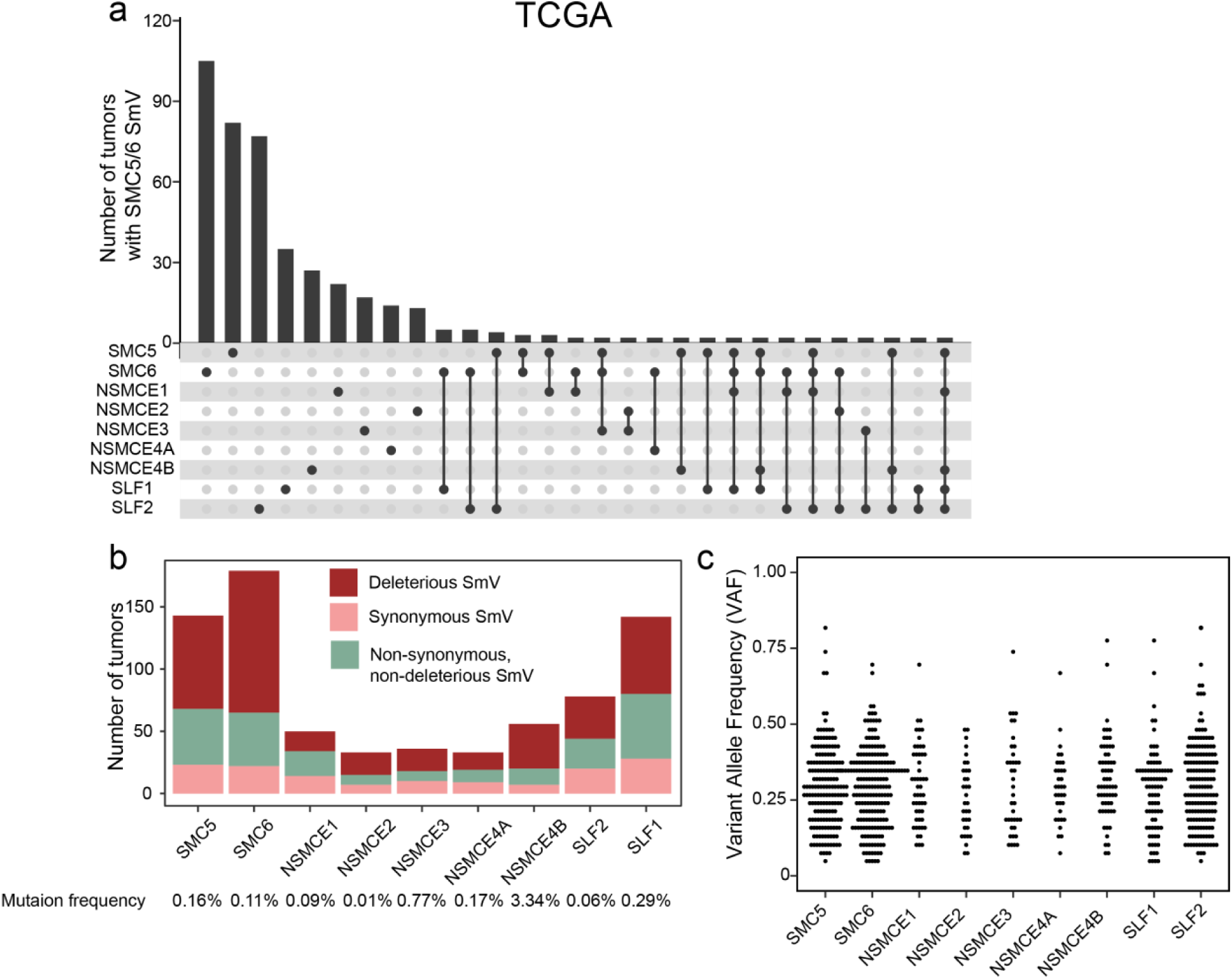
Tumor-associated SmV occur across all SMC5/6 complex genes. (**a**) UpSet plot depicting frequency of SMC5/6 SmV in tumors from TCGA. (**b**) Type and number of SmV in each SMC5/6 gene found in TCGA exomes. Mutation frequency reflects the number of unique SmV relative to size of each gene. (**c**) Variant allele frequency (VAF) of SMC5/6 SmV in TCGA exomes. Each dot represents VAF of the indicated gene from an individual tumor.

**Supplemental Figure 2.**
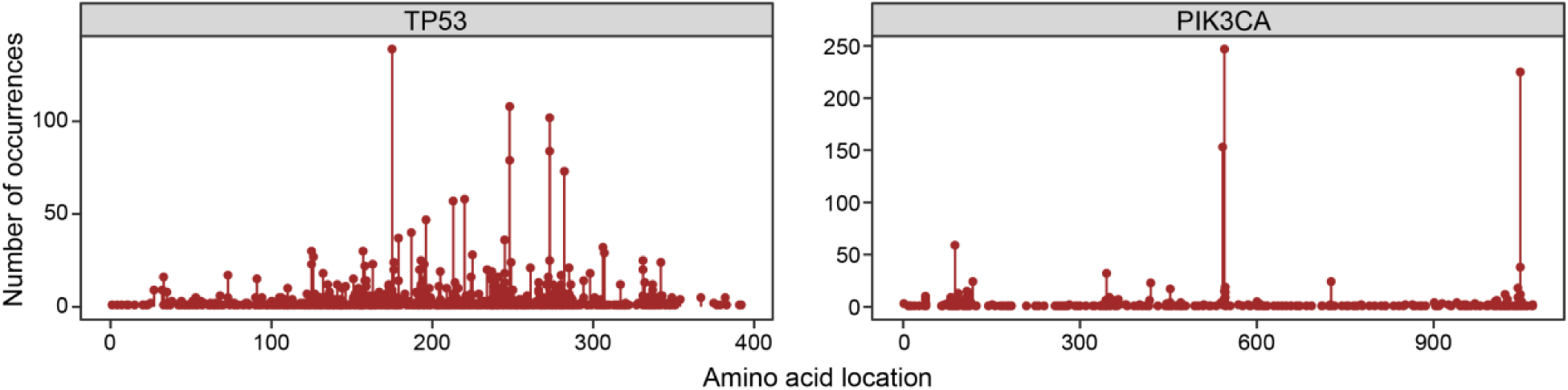
SmV in tumor suppressors and oncogenes occur at hotspots. Lollipop charts depicting frequency of deleterious TP53 and PIK3CA SmV identified in TCGA.

**Supplemental Figure 3.**
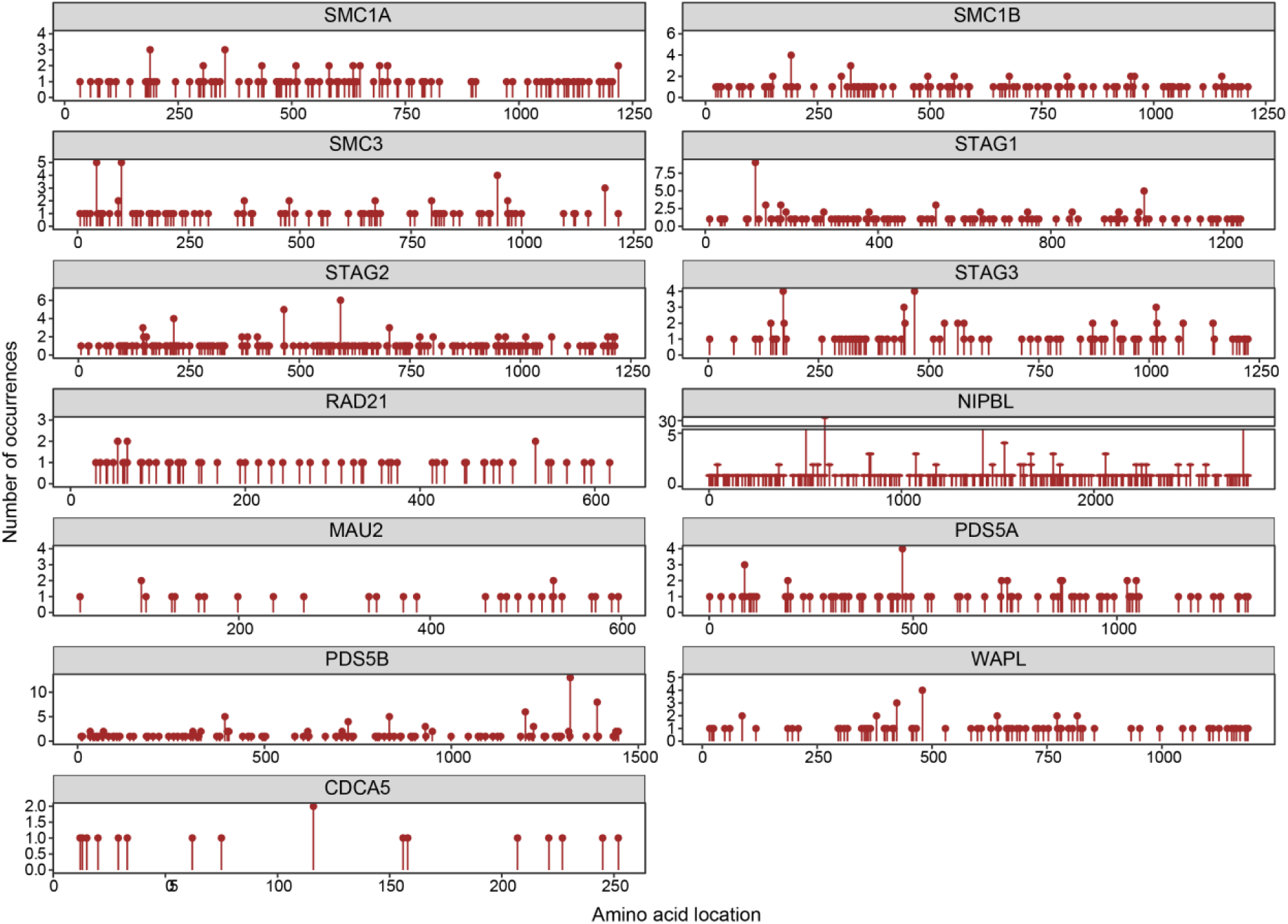
Spectra of cohesin SmV in cancer. Lollipop charts depicting frequency of deleterious SmV found in cohesin complex genes in TCGA exomes.

**Supplemental Figure 4.**
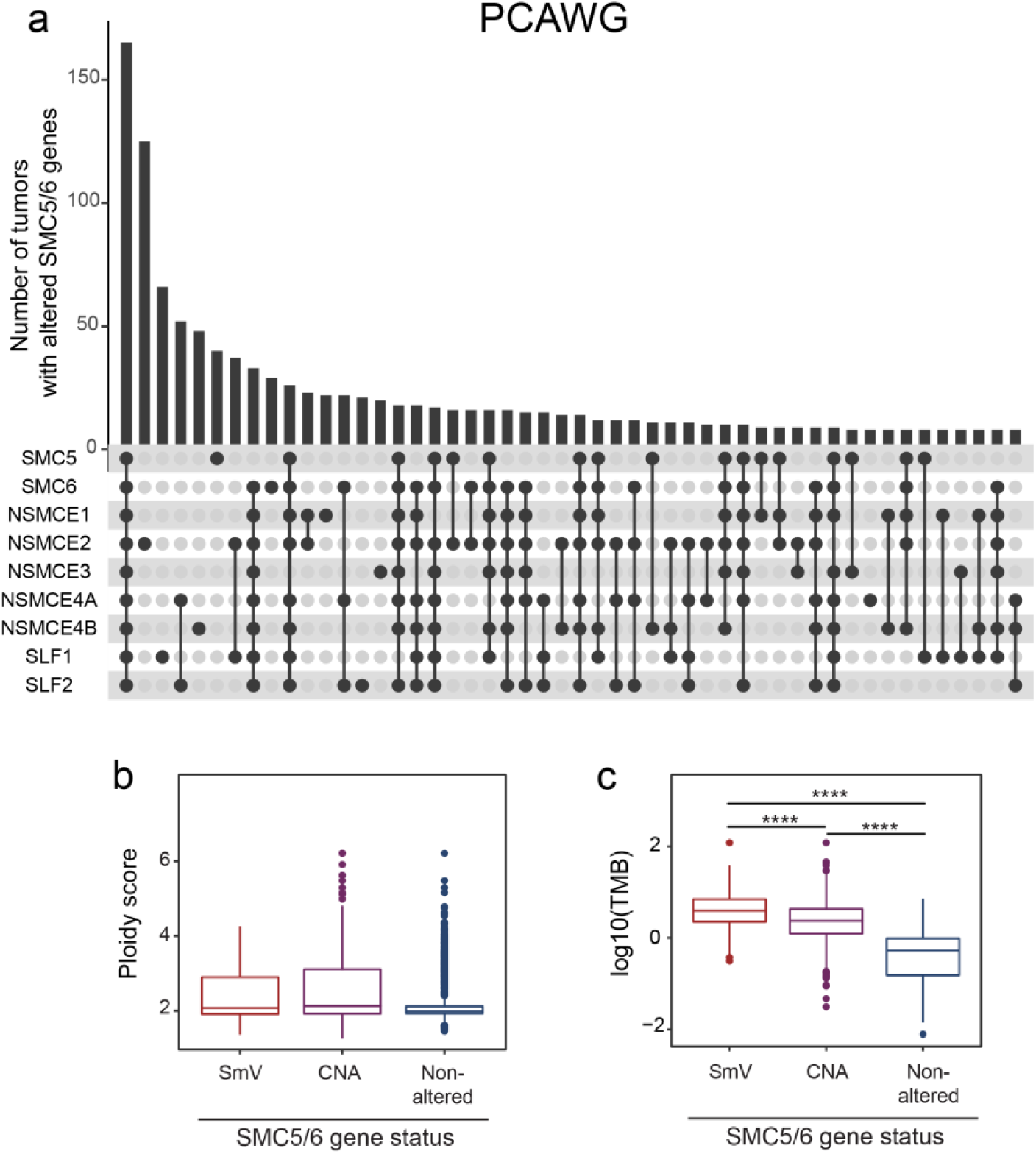
Elevated TMB associated with deleterious SmV in SMC5/6 genes in PCAWG genomes. (**a**) UpSet plot depicting frequency of SMC5/6 alterations (exonic SmV, low, and high CNV) in tumors from PCAWG. (**b**) Ploidy score indicating degree of aneuploidy (median values range 1.988-2.127) and (**c**) TMB in PCAWG cohorts of genomes with SmV, CNA, or non-altered SMC5/6 genes. Significance determined by Wilconxon rank-sum test; **** p<0.0001.

**Supplemental Figure 5.**
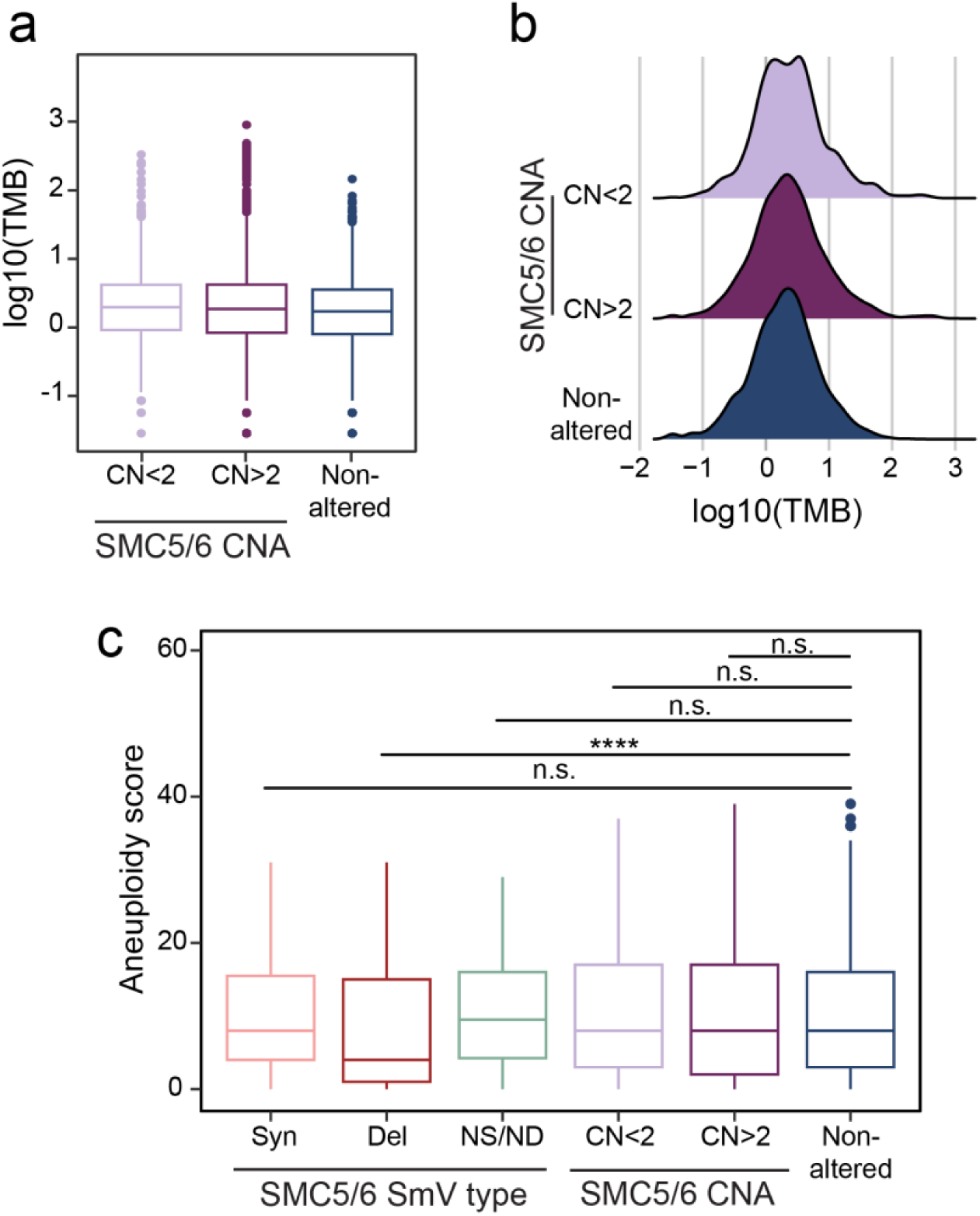
SMC5/6 SmV do not impact tumor aneuploidy. TMB in cohorts of TCGA tumors with copy number alterations in SMC5/6 genes shown as (**a**) box and whisker plot, and (**b**) ridge line plot demonstrating similar TMB (median range 0.3-0.36) between the compared groups. (**c**) Aneuploidy score of TCGA tumor cohorts categorized by type of SmV or copy number alteration in SMC5/6 genes. Significance determined by Wilcoxon rank-sum test; **** p<0.0001.

**Supplemental Figure 6.**
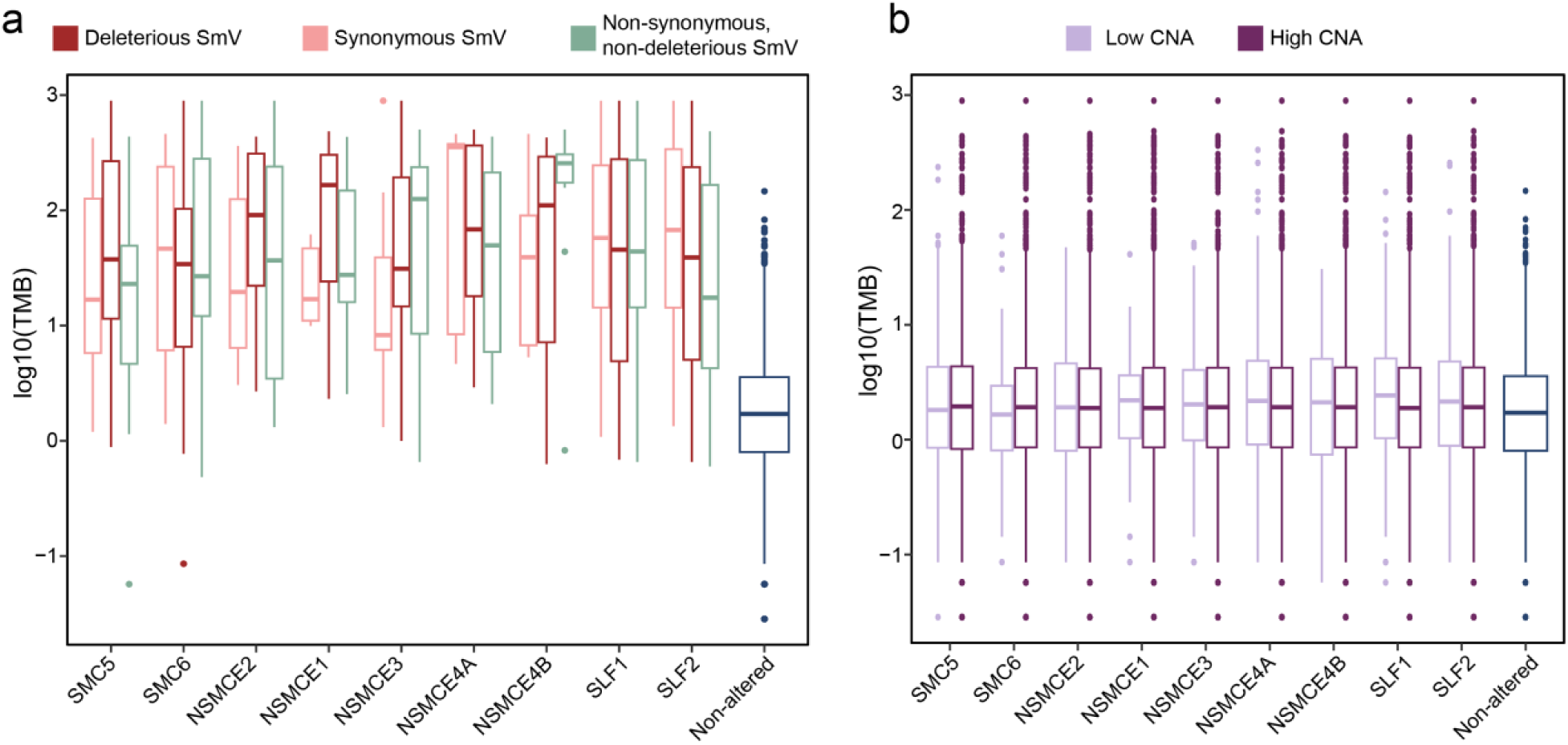
Variants in all SMC5/6 genes are associated with increased TMB. TMB in cohorts of TCGA tumors separated by type of SmV (**a**) or degree of copy number alteration (**b**) in each SMC5/6 gene.

**Supplemental Figure 7.**
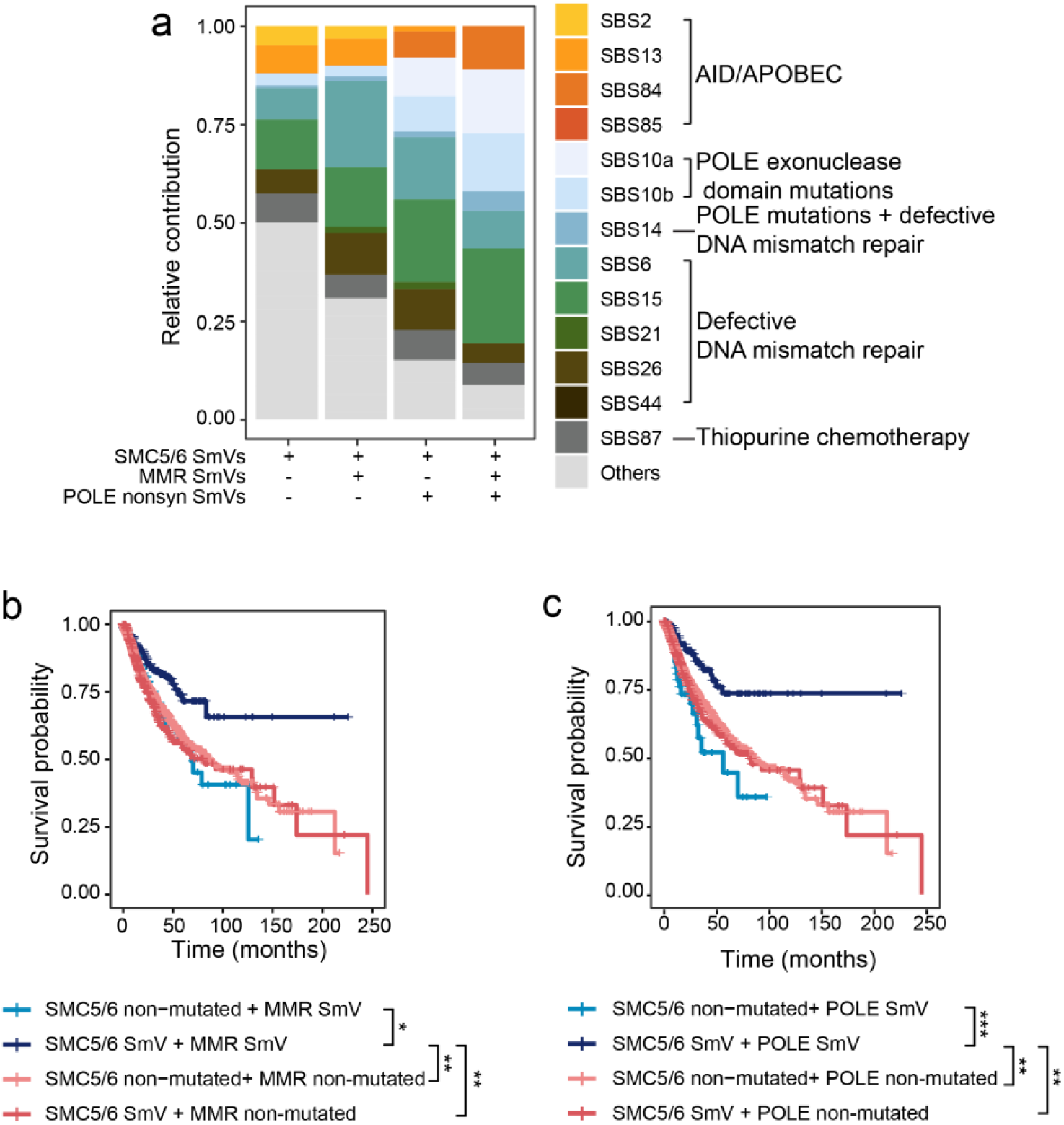
SMC5/6 SmV in combination with POLE or MMR SmV augment survival outcomes. (**a**) Relative contribution of single base substitution (SBS) signatures from TCGA exomes categorized by nonsynonymous SmV in SMC5/6 complex genes and/or MMR genes and POLE. Etiology of SBS signatures is shown in legend. Colored signatures are designated with etiology in legend; these signatures comprised >5% relative contribution of TMB in SMC5/6 deleterious SmV cohort. Signatures in gray are designated “others” and comprised <5% relative contribution of TMB in deleterious SmV cohort (**b-c**) Kaplan-Meier curve of TCGA exomes with non-synonymous SmV in SMC5/6 genes, MMR genes, and/or POLE. p-value determined by log-rank test.

## Notes

### Competing Interest Statement

The authors have declared no competing interest.

### Author Declarations

Somatic variant calls and copy number information of the TCGA projects were obtained using R package TCGA biolinks (version 2.32.0), while whole genome sequencing data of the ICGC was downloaded at https://dcc.icgc.org/releases/PCAWG (release 28). Clinical information, aneuploidy and ploidy scores for both TCGA and PCAWG (ICGC) dataset were obtained from cBioPortal on January 15th, 2024. Sample and patient identification numbers were utilized to match data across these databases. Somatic variant calling and clinical data of colorectal metastases were performed and reported by the Hartwig Medical Foundation (study number NCT01855477). The datasets use only aggregated/summary data. No individual-level data is displayed.

